# Mandated Bacillus Calmette-Guérin (BCG) vaccination predicts flattened curves for the spread of COVID-19

**DOI:** 10.1101/2020.04.05.20054163

**Authors:** Martha K. Berg, Qinggang Yu, Cristina E. Salvador, Irene Melani, Shinobu Kitayama

## Abstract

BCG vaccination may reduce the risk of a range of infectious diseases, and, if so, could serve as a protective factor against COVID-19. Here, we compared countries that mandated BCG vaccination at least until 2000 with to countries that did not (140 countries in total). To minimize any systematic effects of reporting biases, we analyzed the rate of the day-by-day increase in both confirmed cases and deaths in the first 30-day period of country-wise outbreaks. The 30-day window was adjusted to begin at the country-wise onset of the pandemic. Linear mixed models revealed a significant effect of mandated BCG policies on the growth rate of both cases and deaths after controlling for median age, gross domestic product per capita, population density, population size, net migration rate, and various cultural dimensions (e.g., individualism and the tightness vs. looseness of social norms). Our analysis suggests that mandated BCG vaccination can be effective in the fight against COVID-19.

**Teaser:** National policies for universal BCG vaccination are associated with flattened growth of country-wise COVID-19 cases and deaths.

## Introduction

The current pandemic of COVID-19 began in December 2019 in Wuhan, China. Since then, it has rapidly spread across the globe. Currently, there is no end in sight. The present work is motivated by prior evidence that Bacillus Calmette-Guérin (BCG) vaccination (typically given at birth and/or during childhood) offers a long-lasting protective effect not only against tuberculosis (the intended target of BCG), but also against various other infectious diseases (*1-3*). Recent suggestions abound that BCG could be an effective tool in fighting against COVID-19. However, existing cross-national analyses are hampered by methodological weaknesses. For the most part, no effort has been made to exclude potential effects of reporting biases. The potential benefit of universal BCG policies requires careful assessment. To address this gap, we focused on the *rate of the increase* in both confirmed cases and deaths during an early period of country-wise outbreaks, and tested whether this rate might be slower in countries that mandated BCG vaccination at least until 2000, compared to those that did not.

The BCG vaccine is used against tuberculosis (*4*). One review has found that BCG vaccination reduces the risk of tuberculosis by 50% (*5*). A follow-up of an earlier BCG clinical trial performed on native Americans show that BCG protects people from both tuberculosis and lung cancer for up to several decades, throughout each person’s life (*2, 3*). A more recent meta-analysis of a broader range of observational studies and clinical trials (*1*) suggests that the effectiveness of BCG could extend to all-cause mortality. Several controlled trials provide consistent results, showing that the reduced mortality is attributable to protection against respiratory infections, as well as neonatal sepsis (*6-8*). Altogether, the available evidence suggests that BCG has beneficial effects on immunity against a range of lung-related infections that go beyond tuberculosis, which makes it a promising candidate for defending against COVID-19. As for mechanisms, recent experimental work (*9*) finds that BCG vaccination causes genome-wide epigenetic reprogramming of human monocytes, which in turn predicts protection against experimental viral infection.

Over the last century, many countries adopted universal policies of mandatory BCG vaccination to fight against tuberculosis, which was then a major threat. Since then, many countries maintained such a policy at least until very recently (e.g., China, Ireland, Finland, and France). Some other countries terminated the policies as tuberculosis ceased to be a threat (e.g., Australia, Spain, Ecuador). Of note, some countries never mandated BCG vaccination (e.g., U.S., Italy, and Lebanon). Therefore, there is sufficient variability in the presence or absence of such policies distributed across different regions of the world, which makes it possible to draw a systematic comparison.

We examined the day-by-day increase of both confirmed cases and deaths and compared the rate of increase between countries that had mandated BCG policies at least until recently and those that did not. The start of the growth curves was set to be equal across countries, thereby controlling for the varying onset of the pandemic in different countries. Specifically, we focused on a time period either after the first 100 confirmed cases (as in *10)* or after one confirmed COVID-caused death. We then tested the initial, exponential spread of the virus. To exclude any systematic influences of cross-national variation in reporting biases, we focused on the rate of increase of both cases and deaths. These rates are uncontaminated by reporting biases as long as the biases are stable during the period tested. Thus, to avoid any systematic variations in reporting biases, it is important to examine a short initial period of growth. At the same time, it is necessary to test a sufficiently long period to obtain reliable estimates of the growth rate. To simultaneously meet these two competing demands, we chose to examine the first 30 days of the onset of country-wise outbreaks in the main analysis, which was followed by a robustness check testing an even shorter 15-day period. In addition, in a subsequent analysis, we adopted a measure of country-wise reporting biases and weighted the data accordingly. Further, we also controlled for test availability.

We first tested whether the growth rate would be significantly slower in countries that have continued to mandate BCG vaccination at least until the year 2000, as compared to countries that do not currently require it. This year (2000) was chosen since vaccination may become effective at the population level only when a vast majority (70-80% according to a simulation reported in *11*) is made resistant against a target virus, a phenomenon known as "herd immunity” (*12*). In the countries that had mandated BCG at birth at least until the year 2000, a vary majority of adults must have been made resistant against lung-related viral infections. We also explored whether there might be any difference between those that never had such a policy and those that had one during the 20th century but have since terminated the policy for at least a few decades. As a final robustness check, we tested whether the groups of countries that vary in BCG policy status might also vary on various cultural dimensions, such as individualism vs. collectivism (*13, 14*) and the tightness vs. looseness of social norms (*13*).

## Results

### Confirmed Cases

All countries that had reported at least 15 days of at least 100 total confirmed cases, and that had available data on BCG policy and covariates (median age, gross domestic product per capita, population density, population size, and net migration rate) were included (134 countries in total). For each country, day 1 was set to be the first day of at least 100 confirmed cases. See Column 2 of Table S1 for the date of day 1 for each included country.

To model exponential growth of confirmed cases, we estimated a linear mixed model of the natural log-transformed number of confirmed cases. We entered two contrasts designating BCG policy status (current vs. [past and none] combined and past vs. none). The effect of BCG policy status on growth rate is reflected by the interactions between day and each BCG policy status contrast.

As shown in Table 1-A, we found a significant main effect of day, *b* = 0.117, *p* < .001, reflecting an exponential increase in cases over time. This increase was qualified by a significant interaction between day and BCG policy status. Specifically, the growth rate of COVID-19 cases was significantly slower in countries with mandated BCG vaccinations, compared to countries without mandated BCG vaccinations, *b* = −0.041, *p* < .001 (see Fig. 1-A and B). Fig. 2-A shows the distribution of the country-wise regression coefficients.

**Table 1.**
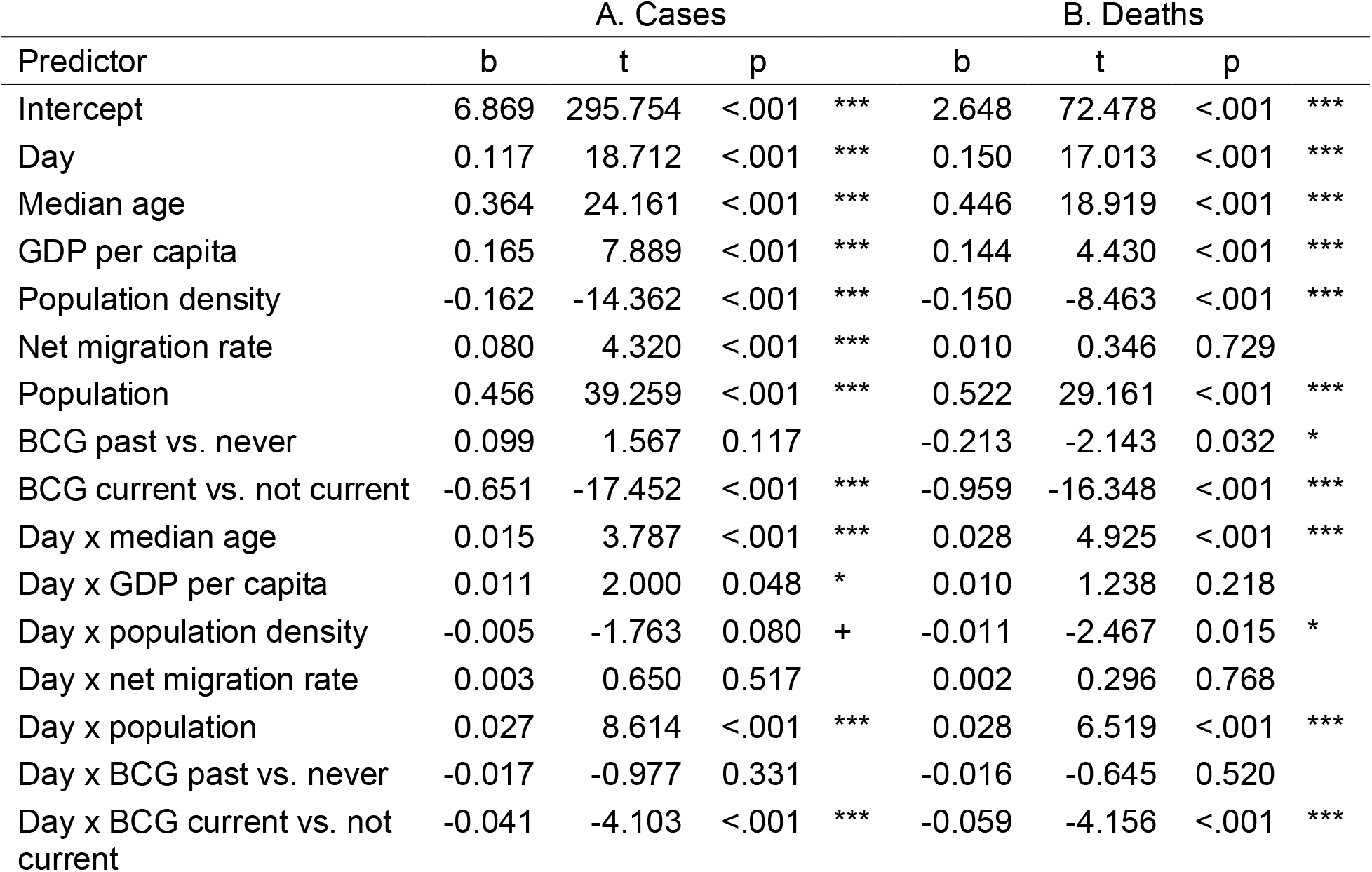
Regression tables predicting growth in (A) cases and (B) deaths. Day is mean centered, and BCG policy variables are both contrast-coded. Population is natural log-transformed, and all covariates are standardized.

**Figure 1.**
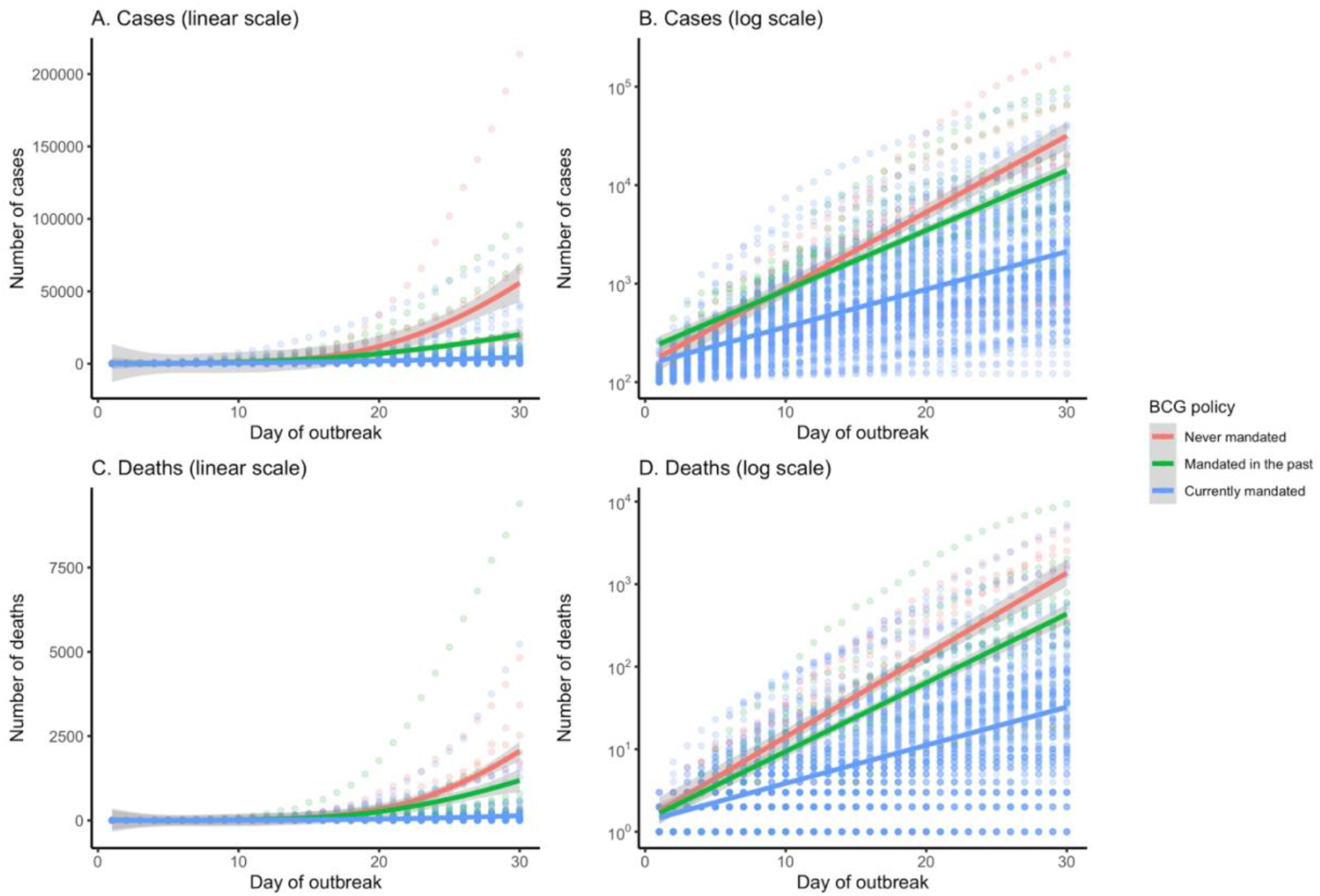
Growth curves by country BCG policy for (A-B) cases and (C-D) deaths, presented on linear (A & C) and logarithmic (B & D) scales.

**Figure 2.**
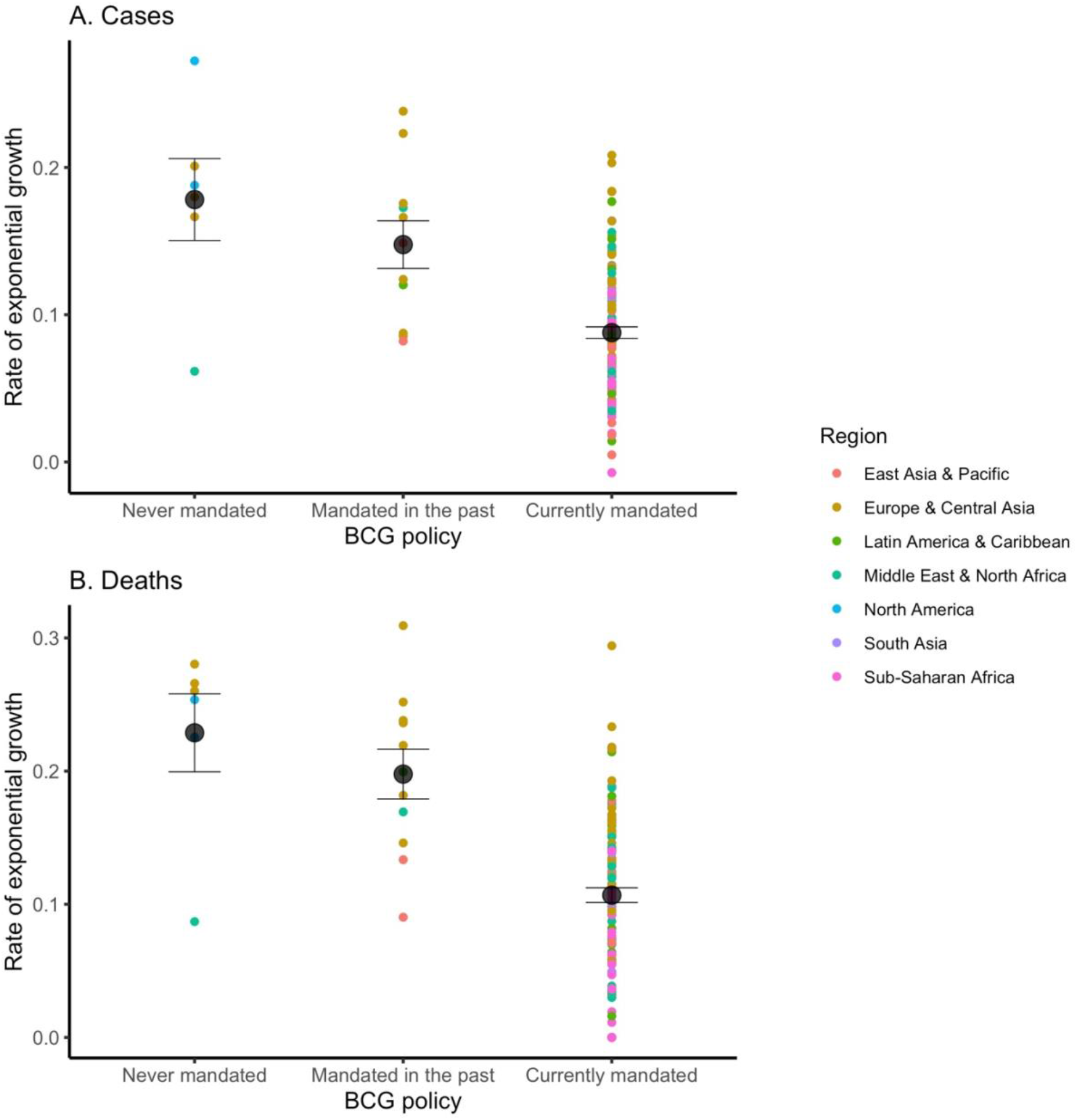
Growth rate of (A) cases and (B) deaths for each country, plotted by BCG policy and region. Growth rate is adjusted by median age, GDP per capita, population density, total population (log-transformed), and net migration rate. Means and standard error are plotted for each group.

Countries that once had such policies but terminated them before 2000 were not significantly different in growth rate from those that never instituted mandatory BCG vaccinations, *b* = −0.017, *p* = .331. In terms of control variables, larger population size, higher GDP per capita, and higher median age predicted a faster growth rate of confirmed cases. See Table S2 for a correlation table of all predictor variables.

The effect of BCG policy status on COVID-19 cases remained unchanged when countries were weighted by reporting quality (see Supplementary Results 1) and when controlling for the total number of tests (see Supplementary Results 2-A). Hence, biases in testing and reporting, demonstrably pervasive across countries, had little or no effect on the effect of universal BCG policies on the growth rate. Moreover, this effect also did not change when a 15-day time window was used (see Supplementary Results 3-A), adding further evidence that the main analysis is unlikely to be due to any systematic variations in reporting biases during the 30-day period. In addition, the BCG effect had little to do with the cultural dimensions of individualism vs. collectivism, power distance, and tightness/looseness (see Supplementary Results 4-A).

### Deaths

All countries that had reported at least 15 days of at least one death from COVID-19, and that had available data on BCG policy and covariates (135 countries in total) were included in this analysis. For each country, day 1 was set to be the first day of at least 1 confirmed death. See column 3 of Table S1 for the date of day 1 for each included country.

We estimated a linear mixed model of the natural log-transformed number of deaths, controlling for the same control variables as above. As in the analysis on confirmed cases, we found a significant main effect of day, *b* = 0.150, *p* < .001, reflecting an exponential increase in deaths over time (Table 1-B). This increase was qualified by a significant interaction between day and BCG policy status. Specifically, the growth rate of COVID-19 related deaths was significantly less in countries with mandated BCG vaccinations, compared to countries without mandated BCG vaccinations, *b* = −0.059, *p* < .001 (Fig. 1-C and D). Fig. 2-B shows the distribution of the country-wise regression coefficients.

Countries that once had such policies but terminated them before 2000 were no different in growth rate from those that never instituted mandatory BCG, *b* = −0.016, *p* = .520. In terms of control variables, larger population size and higher median age predicted a faster growth rate of COVID-19 deaths. Greater population density predicted a slower growth rate of deaths; however, this finding should be interpreted with caution given that the spread of COVID-19 is positively correlated with population density at a more fine-grained level of analysis (e.g., cities), rather than the averaged density at the country level (*14*).

The effect of BCG policy status on COVID-19 related deaths remained unchanged when controlling for the total number of tests (see Supplementary Results 2-B), and when a 15-day time window was used (see Supplementary Results 3-B), showing the robustness of the main analysis. In addition, the BCG effect was unrelated to the cultural dimensions mentioned above (Supplementary Results 4-B).

## Discussion

Our analysis shows that mandatory BCG vaccination is associated with a flattening of the curve in the spread of COVID-19. The effect we demonstrate is quite substantial. For example, our model estimates that the total number of COVID-19 related deaths in the US as of March 29, 2020 would have been 661—27% of the actual figure (2467)—if the US had instituted the mandatory BCG vaccination several decades earlier (see Supplementary Results 5).

Our study is not the first to test the hypothesis that the country-wise spread of COVID-19 might depend on each country’s BCG policy status. However, existing analyses are hampered by their focus on the cumulative totals of confirmed cases and deaths (*15-29*). These tallies depend on how earlier or sooner the onset of the pandemic was in each country. Moreover, they are massively influenced by reporting biases (including the availability of diagnostic testing), which can be both sizable and variable across countries. The same reservation applies to fatality rate (total deaths/total cases) (*18, 26, 28–32*) since the reporting biases are far more likely and cross-nationally variable for the confirmed cases than for the deaths. We circumvented these problems in three ways. First, we focused on the rate of growth of both cases and deaths, which should be uninfluenced by reporting biases as long as these biases are stable during the period of study. To meet this requirement, we focused on a short period (either the first 30 days or 15 days). Second, we used the best available estimate of country-wise reporting biases and used this as a weight in our analysis. Third, we controlled for testing availability.

Notably, the growth curves were as steep in countries that mandated BCG policies only during the 20th century as in those that never mandated the vaccine. BCG vaccination may become effective only when a substantial proportion of the population is made resistant to a virus. That is to say, the spread of the virus may be slowed only when there is "herd immunity” that prevents the virus from spreading easily across the population (see a simulation in (*11*)). Note that as long as others receive vaccination, any single individual will be protected without vaccination, leading to a temptation for free-riding (i.e., not getting vaccinated). Hence, in the absence of state-imposed mandatory vaccination, cultural norms emphasizing prosocial interdependent orientations (*34, 35*) may prove to be crucial for the success of BCG in preventing future outbreaks of COVID-19 (*11, 36*). While the current analysis provided no evidence, this possibility must be addressed in future work.

Some limitations of our effort must be acknowledged. In all national policies, BCG is given early in life, typically at birth. It remains unclear whether BCG vaccination might be effective when given to adults. Nor is it known how long BCG vaccination might provide immunity to COVID-19 although it is effective against tuberculosis and lung cancer for several decades (*2, 3*). Moreover, it is uncertain whether BCG might have any adverse effects when given to those already infected with COVID-19. There is an urgent need for randomized clinical trials. Lastly, the rates of exponential growth showed substantial variability across countries that have mandated BCG vaccination (Fig. 2-A and B). Hence, BCG is by no means a magic bullet that assures safety against COVID-19. In all likelihood, there are some societal variables that moderate this effect. This variation must be addressed in future work.

All these limitations notwithstanding, the current evidence is the first to show a significant advantage of universal BCG policies in reducing the spread of COVID-19, thereby justifying a thorough investigation of the merit of the mandatory BCG vaccination in the fight against COVID-19.

## Methods

### Data

#### Main variables

We retrieved data on daily confirmed COVID-19 cases and deaths by country from a public repository updated daily by the Johns Hopkins University Center for Systems Science and Engineering (https://github.com/CSSEGISandData/COVID-19). Our current results are based on data through June 10, 2020. For confirmed cases, we included countries with at least 15 days of data, starting with at least 100 reported cases as ‘day 1’. For deaths, we included countries with at least 15 days of data, starting with at least 1 reported death as ‘day 1.’

BCG vaccination policy data for each country were compiled from the BCG World Atlas (http://www.bcgatlas.org/index.php) (*37*). Countries were excluded if policy information was unavailable. Data included BCG policy status (vaccination never mandated, vaccination mandated in the past but terminated before 2000, vaccination mandated either currently or up until at least 2000), and, for countries that currently mandate BCG vaccination, the year that the current policy was implemented. We defined this variable based on data from the year 2000, so that ‘vaccination currently mandated’ refers to any country that continued to mandate the BCG vaccination into the 21st century. We created 2 contrast-coded variables to capture BCG policy. The first was a contrast between countries that currently mandate BCG (including those that maintained mandated BCG until at least 2000) and countries that do not currently mandate BCG (including those that terminated mandated BCG before 2000). The second was a contrast between countries that previously mandated BCG that terminated it before 2000 and countries that never mandated BCG.

The 139 included countries are listed in Table S1, which shows the date of the first 100 confirmed cases, the date of the first confirmed death, and the BCG policy status for each of the countries.

#### Demographics

Total population (in thousands) was included since the number of both confirmed cases and deaths should be larger for more populous countries. It was compiled from the United Nations Department of Economic and Social Affairs World Urbanization Prospects 2018 (*38*). Population was natural log-transformed to reduce skewness. Median age of the total population (in years) was included since older adults are more susceptible to viral threats. Population density (in persons per square kilometer) was used because it is likely to foster greater social contact, resulting in greater chances of infection. Net migration (persons entering country minus persons exiting country, per 1000 population) was included so as to control for population movement. These statistics were compiled from the United Nations Department of Economic and Social Affairs World Population Prospects 2019 (*39*). Gross domestic product (at purchasing power parity) per capita (GDP per capita), compiled from the World Bank International Comparison Program database (*40*), was included to control for economic development.

#### Underreporting of cases

Countries may vary in underreporting of COVID-19 cases due to governmental information suppression, a lack of tests, or both. As noted, this variable is likely relatively stable over the 30-day period under study, and therefore, it is unlikely to have systematic influences on the slope of the growth curves in the present analysis. Nevertheless, underreporting may decrease data quality and therefore may cause more subtle biases in the estimation of the slopes. To account for this, we ran the same models and weighted each country based on the accuracy of their reporting.

We used an index of underreporting devised by Russell and colleagues (*41*). Using country-wise data on the estimated likelihood of fatality given the admission to hospitals (1.4%, based on large-scales studies in China and Korea), the researchers have calculated the expected number of patients admitted to the hospitals (the reported number of deaths/.014). They then compared this expected number (cases_expected_) with the reported number of cases admitted to the hospitals (cases_actual_). The underreporting index is the percentage of cases that are unaccounted for (= [cases_expected_ - cases_actual_]/ cases_actual_). Russell et al. (2020) further account for the delay between the admission to hospitals and eventual deaths (estimated based on the distribution of the delay from hospitalization-to-death for cases that are fatal) to improve the accuracy of the estimated underreporting. Some countries, such as Norway, Israel, and South Korea, show substantial underreporting (underreporting index > 50%), whereas some others, such as Italy, Spain, and Morocco, show very low underreporting (underreporting index < 10%). We used country-wise underreporting scores (https://github.com/thimotei/CFR_calculation). Since only daily estimates are available, rather than averages over time, we used estimates from April 15, 2020, which is included in the majority of countries’ 30-day period of data. Due to the lack of available data for some nations, the number of countries included in the analysis of cases dropped from 118 to 77.

#### Number of tests

Countries may vary in the number of COVID-19 tests that are available, which may influence the number of cases and deaths that are reported. As noted, this variable is likely relatively stable over the 30-day period under study, and therefore, it is unlikely to have systematic influences on the slope of the growth curves in the present analysis. Nevertheless, to account for the possibility that our results are explained by differences in testing availability, we ran the same models and controlled for the total number of tests in each country.

We used country-wise numbers of total COVID-19 tests (https://github.com/owid/covid-19-data/tree/master/public/data/) (*10*). Consistent with our underreporting analysis, we used estimates from April 15, 2020. Due to the lack of available data for some nations, the number of countries included in the analysis of cases dropped from 118 to 77.

#### Cultural dimensions

Three cultural dimensions were tested as potential confounding variables. To examine whether BCG policy status might be confounded with some key cultural dimensions, we tested three cultural dimensions. We included individualism vs. collectivism (*42*) and power distance (*43*) since Western individualistic and/or more egalitarian societies tend to have no current mandated BCG policies. The culture scores for the two dimensions were obtained from (*43*). We also included the tightness vs. looseness of social norms (*13*), as this dimension could increase either norm abidance or the rigidity of social systems. Country-wise scores were obtained from (*13*).

### Statistical Analysis

All analyses were conducted on up to 30 days of data from each eligible country. Linear mixed effect models with restricted maximum likelihood estimation were used to analyze both the number of cases and deaths. We first natural log-transformed both cases and deaths in order to account for the exponential nature of the increase of both (*44*). Each model estimated a random intercept, and a random slope across days for each country, to allow for heterogeneity in growth curves between countries. Since our maximal model did not converge, we dropped random intercept from the model. Day was centered so that main effects could be interpreted as differences at the mean day of the growth curve. Models included day, BCG status (with 2 contrasts), and the interaction between day and BCG contrasts. All demographic variables were included along with their interactions with day: median age, population density, net migration, total population and GDP per capita. Total population was natural log-transformed to reduce skewness. All demographic and cultural variables were standardized.

## Data Availability

All data is available in the public domain.

